# Poverty and family adversity trajectories and Not in Education, Employment or Training (NEET) status in early adulthood: Evidence from the UK Millennium Cohort Study

**DOI:** 10.1101/2025.10.07.25337496

**Authors:** Kalu Udu, Nicholas Kofi Adjei, Akanni Lateef, Gianmaria Niccodemi, Yanhua Chen, Yu Wei Chua, Rosalie Cattermole, Michelle Black, Luke Munford, Karsten Thielen, Leonie K Elsenburg, Naja Hulvej Rod, Steven Hope, Hanna Creese, Dougal Hargreaves, David C Taylor-Robinson

## Abstract

**Background:** Young people who are not in education, employment or training (NEET) are at an increased risk of long-term social and economic disadvantage. While previous research has linked various risk factors and individual characteristics to NEET status, evidence on the cumulative impact of early-life exposure to childhood adversity in the UK remains limited.

**Methods:** We analysed longitudinal data on 8,368 participants from the UK Millennium Cohort Study. Using group-based multi-trajectory modelling approach, we identified six distinct exposure trajectories of poverty and family adversity (including poor parental mental health, domestic violence and abuse and alcohol use) from aged 9 months to 14 years. NEET status was assessed at age 17. Adjusted odds ratios (aORs) and 95% CIs were estimated using logistic regression models. Population attributable fractions (PAFs) were calculated to estimate the proportion of NEET cases attributable to childhood poverty and family adversity.

**Results:** Overall, 3.5% of participants were NEET at age 17 years. NEET status was more prevalent among young people from socially disadvantaged backgrounds than their peers. Exposure to persistent family childhood adversities was associated with greater likelihood of being NEET. Young people exposed to both persistent poverty and poor parental mental health throughout childhood (0-14 years) had five times greater odds of being NEET (adjusted odds ratio [aOR] 5.0; 95% CI 3.4–7.5) compared to those in low poverty and adversity. An estimated 52.9% (95% CI: 41.1–61.7) of NEET cases were attributable to persistent exposure to poverty and family adversity.

**Conclusion:** Family childhood adversities, particularly household poverty and poor parental mental health are strongly associated with an increased risk of being NEET on transition to adulthood. Interventions that address early-life socio-economic disadvantage and family functioning may be critical for preventing NEET and mitigating its long-term social and economic consequences.

**What is already known on this topic:**

- Young people who are not in education, employment or training (NEET) are at risk of poor health, social exclusion and long-term economic disadvantage.
- Childhood poverty and family adversity have been associated with NEET status, but their cumulative and life-course impact in the UK remains unclear.

**What this study adds:**

- Using longitudinal data from a nationally representative UK cohort, this study shows that persistent exposure to poverty and family childhood adversity including poor parental mental health increase the likelihood of being NEET at age 17.
- Individuals exposed to multiple family childhood adversity (i.e., poverty and poor parental mental health) were five times more likely to be NEET.
- An estimated 52.9% NEET cases were attributable to persistent poverty and family childhood adversity.

**How this study might affect research, policy, or practice:**

- Interventions that address family childhood adversity, particularly household poverty and poor parental mental health could substantially reduce NEET prevalence and mitigate long-term inequalities.

## Introduction

Young people who are not in education, employment, or training (NEET) are widely recognised as vulnerable population [1, 2], with significant implications for social and public health policy. The concept of NEET has gained considerable attention globally and it is increasingly used by policymakers as a key measure of youth disengagement from the labour market [1, 3, 4]. In the UK, approximately 13.4% of young people aged 16 to 24 were classified as NEET by the end of 2024, marking an increase of over 100,000 compared to the previous year [5].

NEET status has been consistently shown to be associated with a range of adverse outcomes including poor physical health, mental health and early mortality [6–9]. The consequences of non-participation in education, employment or training extend beyond health outcomes [1]. At the societal level, being NEET poses significant social and economic costs, including lost productivity, increased welfare dependency, and employment challenges [3, 10]. Prior evidence also suggests that even brief periods of non-participation from employment, education or training during adolescence may have an enduring ‘scarring’ effect [11], diminishing future employment prospects and reducing future earning potential [12].

Existing literature has identified a range of risk factors for NEET status, including low educational achievement, psychosocial difficulties and substance use [13–20]. More recently, growing attention has focused on the role of early childhood adversities such as poverty, material deprivation, exposure to traumatic events [21] and parental mental illness [22]. These adversities can disrupt normative developmental processes and may further erode the emotional, cognitive and social capacities for a successful transitions into adulthood [23].

Recent Danish population-based registry studies have shown how patterns and trajectories of childhood adversity influence a range of adverse adult outcomes, including NEET. Using the DANLIFE cohort, Elsenburg and colleagues identified that exposure to cumulative adversity trajectories was strongly associated with increased risk of being NEET in early adulthood [22]. In a complementary analysis, Bennetsen et al. [24] showed that early school leaving partially mediated the long-term association between childhood adversity and long-term social benefit, highlighting the role of disrupted education pathways. Further evidence using the DANLIFE cohort demonstrated that individuals exposed to adversities had markedly higher use of health, social and justice services in early adulthood [25], underscoring the broad and intersecting impacts of childhood disadvantage and adversities on public systems.

These studies offer compelling insights into the complexity and clustering of childhood adversities using rich, linked administrative data. However, to date, no analyses have assessed these relationships in the UK context. Building on our previous study on the clustering of poverty and family adversity across the early life course [26], this study extends the evidence base by exploring how trajectories of poverty and family adversity from early childhood through to adolescence are associated with NEET status in early adulthood in a contemporary UK cohort. By adopting a life-course perspective, we aim to capture how poverty and family adversities including poor parental mental health accumulate, persist and interact across key stages of childhood developmental stages to influence NEET status. This approach is crucial to understand how the synergistic effects of multiple childhood adversities shape developmental trajectories and pathways into adulthood [27–29]. We also examine how broader structural indicators of social disadvantage (i.e., household income and maternal education) are linked to NEET status, in order to situate patterns of childhood adversity within the wider context of socioeconomic inequality [30].

## Methods

### Study setting and Participants

We used data from the Millennium Cohort Study (MCS), a large-scale longitudinal study tracking over 18,000 children born in the UK (England, Northern Ireland, Scotland and Wales) between September 2000 and January 2002. The cohort has been followed through multiple survey waves, with data collection occurring when the children were approximately 9 months, 3 years, 5 years, 7 years, 11 years, 14 years and 17 years. The age 17 data were collected from January 2018 to April 2019. The numbers of participants at each wave were 18,552, 15,590, 15,246, 13,857, 13,287, 11,726 and 10,625, respectively. MCS has collected information directly from the main caregiver, usually the child’s mother, and more recently, from cohort members themselves. Additional information on response patterns, sampling methodology and survey design is reported elsewhere [31]. Each data collection wave has received NHS research ethics approval, and the analysis reported here did not require additional ethical approval.

### Outcome and exposures

The main outcome for this study was NEET status at age 17. We derived a binary indicator of NEET status using responses from cohort members to four questions: (1) “Are you currently going to school or college?”, (2) “Are you currently doing an Apprenticeship?”, (3) “Are you currently doing any kind of traineeship, training course or scheme?”, and (4) “Are you currently doing any kind of paid job? Participants who responded “no” to all four items were classified as NEET; those who responded “yes” to at least one were classified as not NEET.

Our primary exposures were trajectories of poverty and family adversity from infancy to early adolescence (9 months to 14 years). Poverty was defined as household equivalised income below 60% of the national median, according to the Organisation for Economic Co-operation and Development household equivalence scale [32]. Family adversity comprised repeated exposure to parental mental health, domestic violence and abuse and alcohol use (see supplementary box S1 for the list and description of measures). In our previous study, we identified six distinct trajectories of poverty and family adversity [26], using group based multi-trajectory modelling approach [28]: (1) the ‘low poverty and adversity’ group (45.1%) includes children with low overall exposure to childhood poverty and family adversity; (2) the ‘persistent poverty’ group (21.0%) includes children with a high probability of experiencing poverty throughout childhood; (3) the ‘persistent poor parental mental health’ group (12.0%) is mainly characterised by high rates of poor parental mental health over time; (4) the ‘persistent parental alcohol use’ group (8.2%) include children with a high probability of being exposed to parental alcohol use throughout childhood; (5) the ‘domestic violence and abuse’ group (3.5%) include children with a high probability of having been exposed to domestic violence throughout childhood; and (6) the ‘persistent poverty and poor parental mental health’ group (10.1%) includes children with high exposure to the co-occurrence of persistent poverty and poor parental mental health over time. These trajectory groups were used in this present study as the main exposure variables [26].

### Confounders

We adjusted for potential confounders associated with childhood adversities in early-life and NEET, guided by a directed acyclic graph (Figure S1). These included child sex, maternal education and ethnicity, all measured when the child was aged 9 months. Maternal ethnicity was categorised as: White, Mixed, Indian, Pakistani and Bangladeshi, Black or Black British, and other ethnic groups. Maternal education was classified as: degree or higher, diploma, A-levels, GCSEs A*–C, GCSEs D–G, or no formal qualifications.

### Statistical Analysis

First, exposure trajectories were characterised from age 9 months to 14 years using group-based multi-trajectory models [26]. This approach identified sub-groups of cohort members who followed similar patterns of adversity from childhood to early adolescence. We then described sample characteristics across the six estimated trajectory groups using percentages (%). In addition, we estimated the prevalence of being NEET at age 17 by maternal education and household income quintile. Differences in prevalence were assessed using Pearson’s χ² tests, with a two-sided threshold of p<0.05. To assess the associations between the trajectory groups and NEET status, we fitted logistic regression models and reported odds ratios (ORs) with 95% confidence intervals (CI), adjusting for potential confounders. The model incorporated longitudinal survey weights to account for non-response, attrition and sampling design. Finally, we estimated population attributable fractions (PAFs) to quantify the proportion of NEET cases that could be potentially prevented if exposure to poverty and family adversity were reduced to the levels observed in the low poverty and adversity trajectory group. PAFs and their corresponding 95% CIs were calculated using the ‘punaf’ command in STATA (see technical description S1 for more details on the model specification). All analyses were conducted using STATA version 16.0.

## Results

### Study Population Characteristics

Of the 10,625 participants at age 17 (wave 7), 8,368 were included in the final analysis (Figure S2). Table 1 presents the baseline characteristics of cohort members and NEET status across the trajectory groups. Estimates derived using multiple imputation by chained equation (n=25) are provided in Table S1. At age 17, 3.5% of cohort members reported being NEET. Those exposed to poverty and family adversity, either in isolation or in combination were generally more likely to be NEET. For example, NEET prevalence was 9.5% among young people in the persistent poverty and poor parental mental health trajectory group compared to 1.6% in the low adversity and poverty group. Differences between the trajectory groups based on socioeconomic status and ethnicity were also evident (Table 1).

**Table 1.** Baseline characteristics and outcome by the six estimated trajectory groups, observed data.

| Characteristics | Family adversity and poverty trajectories |  |  |  |  |  | Overall (N=8368) |
| --- | --- | --- | --- | --- | --- | --- | --- |
|  | Low poverty and adversity (n=3846) | Persistent parental alcohol use (n=706) | Persistent domestic violence and abuse (n=300) | Persistent poor parental mental health (n=991) | Persistent poverty (n=1714) | Persistent poverty and poor parental mental health (n=811) |  |
| **Outcome** |  |  |  |  |  |  |  |
| NEET | 63 (1.6%) | 9 (1.3%) | 10 (3.3%) | 35 (3.5%) | 95 (5.5%) | 77 (9.5%) | 289 (3.5%) |
| No NEET | 3783 (98.4%) | 697 (98.7%) | 290 (96.7%) | 956 (96.5%) | 1619 (95.5%) | 734 (90.5%) | 8079 (96.5%) |
| **Child's sex** |  |  |  |  |  |  |  |
| Boy | 1815 (47.2%) | 319 (45.2%) | 152 (50.7%) | 463 (46.7%) | 707 (41.2%) | 392 (48.3%) | 3848 (45.9%) |
| Girl | 1941 (50.5%) | 361 (51.1%) | 144 (48.0%) | 498 (50.3%) | 923 (53.9%) | 363 (44.8%) | 4230 (50.6%) |
| Missing | 90 (2.3%) | 26 (3.7%) | 4 (1.3%) | 30 (3.0%) | 84 (4.9%) | 56 (6.9%) | 290 (6.5%) |
| **Maternal education** |  |  |  |  |  |  |  |
| Degree plus | 1187 (30.7%) | 309 (43.8%) | 69 (23.0%) | 211 (21.3%) | 43 (2.5%) | 14 (1.7%) | 1833 (21.9%) |
| Diploma | 503 (13.1%) | 81 (11.5%) | 47 (15.7%) | 91 (9.2%) | 55 (3.2%) | 16 (2.0%) | 793 (9.5%) |
| A-levels | 499 (12.9%) | 69 (9.8%) | 40 (13.3%) | 116 (11.7%) | 106 (6.2%) | 35 (4.3%) | 865 (10.3%) |
| GCSE A-C | 1162 (30.2%) | 161 (22.8%) | 88 (29.3%) | 350 (35.3%) | 514 (30.0%) | 233 (28.7%) | 2508 (30.0%) |
| GCSE D-G | 192 (5.0%) | 25 (3.5%) | 27 (9.0%) | 89 (8.9%) | 232 (13.5%) | 114 (14.1%) | 679 (8.1%) |
| None | 212 (5.6%) | 35 (5.0%) | 25 (8.3%) | 103 (10.4%) | 674 (39.3%) | 336 (41.4%) | 1385 (16.6%) |
| Missing | 91 (2.5%) | 26 (3.6%) | 4 (1.4%) | 31 (3.2%) | 90 (5.3%) | 63 (7.8%) | 305 (3.6%) |
| **Maternal ethnicity** |  |  |  |  |  |  |  |
| White | 3442 (89.5%) | 665 (94.2%) | 252 (84.0%) | 821 (82.9%) | 1006 (58.7%) | 465 (57.3%) | 6651 (79.5%) |
| Mixed | 19 (0.5%) | 5 (0.7%) | 6 (2.0%) | 7 (0.7%) | 27 (1.6%) | 16 (2.0%) | 80 (1.0%) |
| Indian | 118 (3.1%) | 2 (0.3%) | 17 (5.7%) | 36 (3.6%) | 52 (3.0%) | 19 (2.3%) | 244 (2.9%) |
| Pakistani and Bangladeshi | 41 (1.1%) | 0 (0.0%) | 5 (1.7%) | 28 (2.8%) | 391 (22.8%) | 193 (23.8%) | 658 (7.9%) |
| Black or Black British | 79 (2.1%) | 3 (0.4%) | 12 (4.0%) | 26 (2.6%) | 110 (6.4%) | 37 (4.6%) | 267 (3.2%) |
| Other ethnic groups | 50 (1.3%) | 4 (0.6%) | 4 (1.3%) | 41 (4.1%) | 39 (2.3%) | 22 (2.7%) | 160 (1.9%) |
| Missing | 97 (2.4%) | 27 (3.8%) | 4 (1.3%) | 32 (3.3%) | 89 (5.2%) | 59 (7.3%) | 308 (3.6%) |

Figure 1a and 1b show the prevalence of NEET status by maternal education level and household income quintile, respectively. NEET status in early adulthood was more prevalent among young people from socially disadvantaged backgrounds. For instance, those born to mothers with lower educational qualifications were more likely to be NEET (Education — degree plus: 0.7% vs. no qualifications: 5.9%). An even steeper social gradient was observed with household income, where NEET prevalence was 7.4% in the lowest income quintile compared to 0.1% in the highest.

**Figure 1a.**
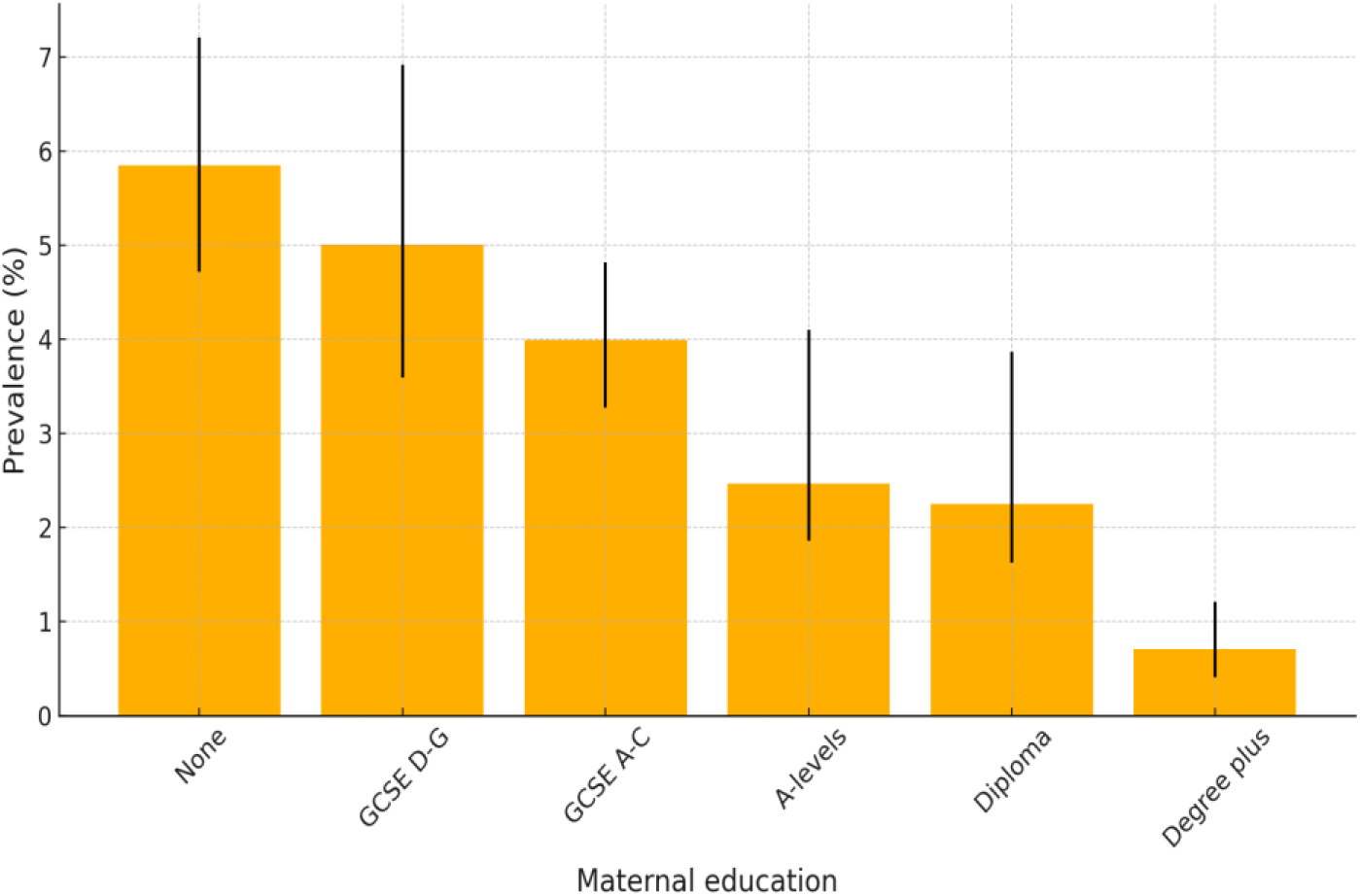
Prevalence (%) and CIs (95% CI) of being NEET in the UK at age 17 by maternal education at birth.

**Figure 1b.**
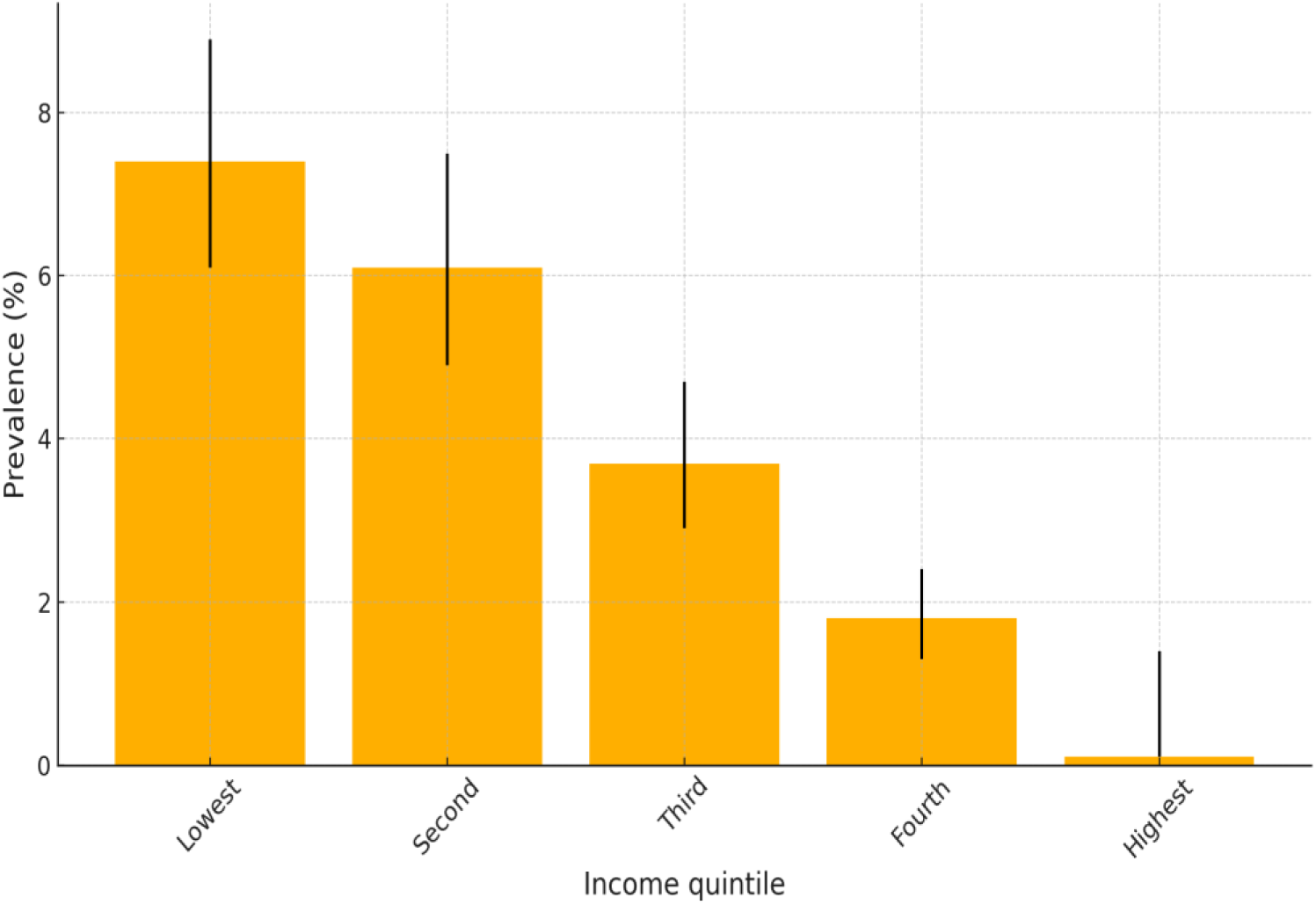
Prevalence (%) and CIs (95% CI) of being NEET in the UK at age 17 by parental income quintile at birth.

### Associations of poverty and family adversity trajectories with NEET status

Figure 2 shows the associations between adversity trajectory group membership and NEET status at age 17. In both the unadjusted model (Table S2) and the adjusted model (Figure 2), participants in trajectories characterised by persistent poverty and family adversity generally had higher odds of being NEET compared to those in the low poverty and adversity trajectory group. Associations were strongest among those exposed to both persistent poverty and poor parental mental health. For example, the odds of being NEET were five times higher among participants in this trajectory group compared to those in the low poverty and adversity group (adjusted odds ratio 5·0; 95% CI 3·4–7·5). The regression analysis was also repeated analysis using imputed data (i.e., multiple imputations by chained equations (n=25)) (Table S3), and the estimates were similar to the main results.

**Figure 2.**
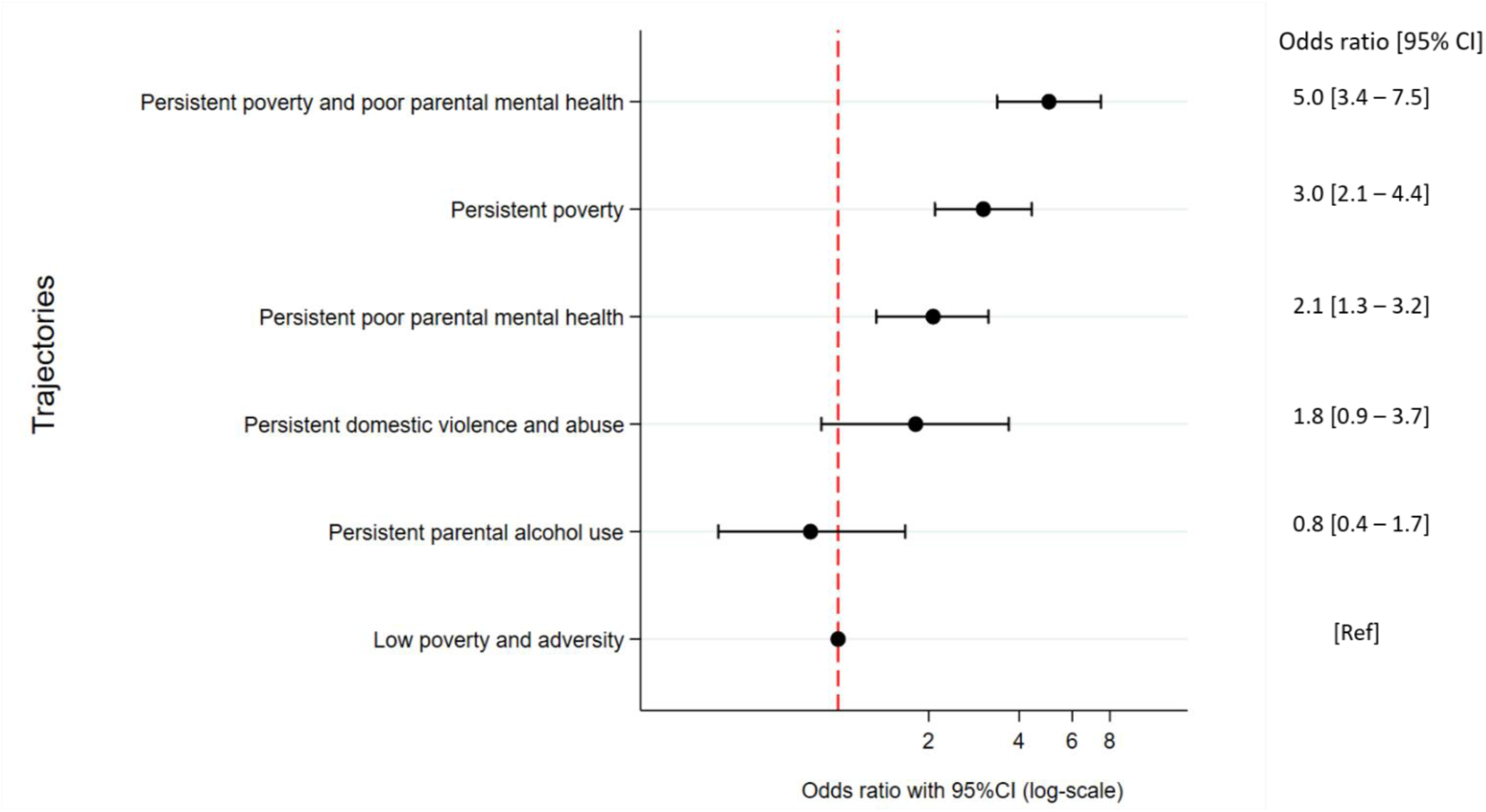
Associations of family adversity and poverty trajectories and being NEET at age 17 years in the UK Millennium Cohort Study. Model adjusted for child’s sex, maternal education and ethnicity.

### Population attributable fraction (PAF)

The PAF estimates show the proportion of NEET cases at age 17 that were attributable to each trajectory group (Figure 3), assuming a causal relationship. Approximately 53% of NEET cases at age 17 were attributable to persistent poverty and family adversity during childhood. This suggests that, if all children had experienced the trajectory characterised by low poverty and family adversity, the number of NEET cases could be reduced by more than half. When broken down into individual trajectory, exposure to persistent poverty alone and in combination with persistent poor parental mental health accounted for the largest proportion (> 50%) of the population-level burden.

**Figure 3.**
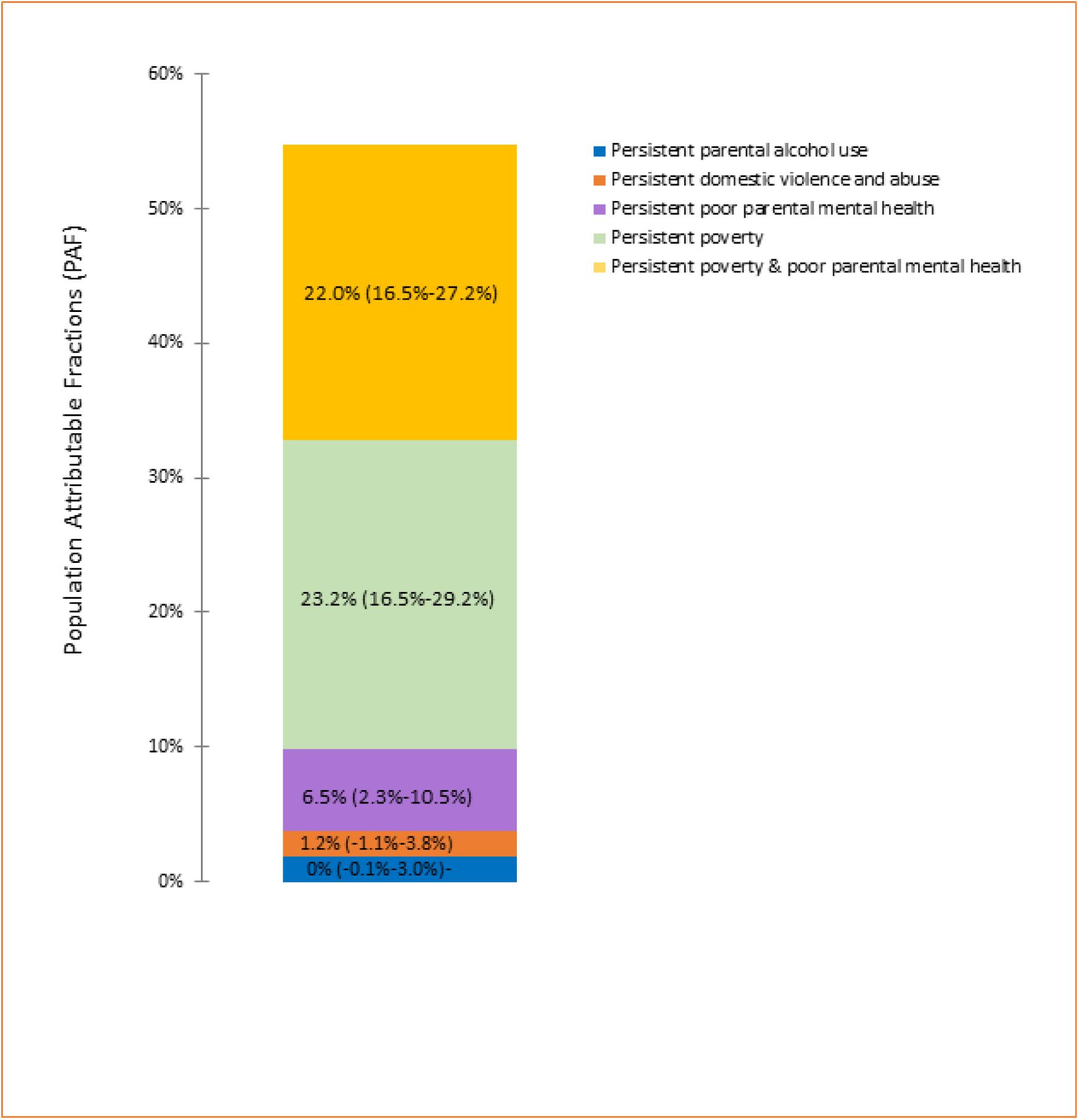
Population-attributable fractions of trajectory groups. Compared to the low poverty and family adversity trajectory groups, the overall proportion of being NEET attributable to persistent poverty and family adversity was 52.9% (95% CI 41.1%–61.7%). Model adjusted for child’s sex, maternal education, and ethnicity

## Discussion

In a large contemporary, population-based cohort from the UK, we show that trajectories of poverty and family adversity across childhood are associated with the risk of being not in education, employment or training (NEET) at age 17. Persistent exposure to poverty, particularly when combined with poor parental mental health was associated with markedly higher risks of NEET status. We found that 3.5% of young people aged 17 reported being NEET, consistent with the national estimate of 3.6% for 16-17 years olds in 2017 [33] — the year participants in the MCS were also around age 17. More importantly, our findings indicate that over half of NEET cases in this cohort were attributable to childhood poverty and family adversity including poor parental mental health.

Our findings are similar to those in the recent Danish register-based study by Elsenburg and colleagues [22], whereby children experiencing high levels of adversity had around 5 times greater risk of being NEET in two consecutive years [22]. Our results are also consistent with prior studies identifying socioeconomic disadvantage, parental unemployment and mental health problems as key risk factors for NEET status [13–16]. The Danish studies and this current analysis highlight the importance of life-course analyses, capturing the clustering and cumulative burden of poverty and other family adversities from infancy through to transition to adulthood [27, 28]. Life-course and trajectory-based approaches have been shown to be useful in modelling how early exposures shape long-term outcomes and how the clustering of multiple adversities can have strong negative effects on young people’s life chances [34, 35].

Indeed, the strength of the associations observed, particularly among those exposed to both persistent poverty and poor parental mental health, suggest a heightened vulnerability [36]. While we did not formally test for interactions over time, the findings are consistent with the possibility that social disadvantage may operate in a cumulative and synergistic manner [26, 35]. The underlying mechanisms are likely to operate through multiple pathways [37]. For example, parental mental health illness may impact children directly through impaired emotional bonding and increased family conflict [36] and indirectly by contributing to poor academic performance and social emotional behavioural problems [38, 39]. These risk factors may form a critical component of the pathway to later NEET status. In fact, academic attainment, school attendance and engagement are known to be key mediators in the pathway between childhood adversity labour market disengagement [1, 40], and these risk factors may further be intensified by reduced access to educational support and structural inequalities [1, 38, 40]. The steep socioeconomic gradient in NEET prevalence marked by higher risk among young people from low-income households and those born to less educated mothers, observed in this study, is consistent with broader literature on the intergenerational transmission of disadvantage [41, 42].

A key strength of this study is the use of a large, nationally representative cohort and the application of a robust trajectory modelling approach to capture patterns of childhood adversity over time [28, 29]. Nonetheless, some limitations should be acknowledged. First, NEET outcome was measured at a single time point and may not reflect temporal patterns or transitions in and out of education or employment [22]. However, as cohort members were aged 17, this outcome measure reflects a critical period immediately following the end of compulsory education. Second, although we adjusted for relevant confounders, residual confounding from unmeasured factors such as school-level factors and peer effects and disabilities may persist [43]. Nonetheless, we assessed the robustness of unmeasured confounding using the E-values approach [44], and we found that our findings are robust to omitted confounding (Figure S3). Third, there are also some limitations related to the use of PAFs. For instance, PAFs rest on the assumption that the observed relationship between exposure and outcome reflects a true causal relationship [45]. However, when interpreted cautiously, PAFs provide a useful tool for estimating the potential population-level impact of modifiable exposures, offering valuable insights for public health planning and policy prioritisation [45]. Fourth, the exposure is limited to poverty and family adversity measures. We lacked data on other dimensions of childhood adversity such as parental psychiatric illness and unemployment, which has been shown to impact later life [35]. Fifth, missing data and attrition are common sources of bias, particularly selection bias, in longitudinal studies. Nonetheless, we addressed part of the missing data using multiple imputation. Finally, our analysis did not directly examine possible mediating factors such as adolescent mental health, academic attainment or school exclusion, which likely lie on the causal pathway. These mechanisms warrant further investigation in future research.

### Policy Implications

Our findings have important implications for both social and public policy. Population attributable fraction (PAF) estimates suggest that, assuming a causal relationship, up to half of NEET cases in this cohort could potentially be prevented if exposure to poverty and family adversity throughout childhood were eliminated. From a public health perspective, addressing early-life poverty and family adversity presents a powerful opportunity to reduce NEET and promote social and economic resilience [46]. Our analysis suggest that this approach could lead to a large reduction in the burden of NEET with huge-cost savings across all sectors. For instance, according to the most recent population estimates, there were around 1,567,000 16-17-year olds in the United Kingdom in 2023 [47]. The figures estimated here suggest that 3.5% of these, around 54,850 16-17-year olds, will be NEET. Applying costs tariffs from in HM Treasury’s Green Book [48] indicate that the average yearly cost to the government, comprising benefit payments and foregone tax and national insurance contributions to be £11,474 per-NEET-per-year. Analysis in this current study shows that an estimated 52.9% (41.1%–61.7%) of NEET cases were attributable to persistent exposure to poverty and family adversity. Therefore, if this were to be removed – i.e. if no children in the UK had the exposure trajectory of low poverty and family adversity – we would see a reduction in NEET by around 1.85% (95% CI: 1.23% to 2.41%) across all 17-year olds, equivalent to around 29,000 (95% CI: 19,300 to 37,700) people. These estimates suggest that the annual saving to the government would be around £332,895,219 (95% CI: £221,690,416 to £432,646,916) per-year.

Indeed, increasing levels of youth NEET and rising social welfare benefit costs related to young people with mental health problems are key policy issues in the UK [49–51]. Our analyses of the MCS population are pertinent to these debates, since MCS captures representative data from childhood on the UK’s young adults who are currently transitioning into the labour market. Our findings highlight the need for a life-course and social determinants prevention perspective. While the UK government has taken steps to expand early years support and has announced plans to address child poverty [52], more action is needed. Priorities should include improved access to parental mental health services, including perinatal and family-based care, anti-poverty strategies such as increasing child benefit, raising the minimum wage, strengthening universal credit protections, stronger support in education, particularly for adolescents at risk of exclusion [53]. These should be coupled with guaranteed training places, apprenticeships and supported employment schemes for vulnerable youth [54]. Without bold investment in upstream interventions, cycles of intergenerational disadvantage will continue to limit opportunities and may further undermine long term economic stability.

## Conclusion

In conclusion, this study provides evidence that persistent exposure to childhood adversity, particularly household poverty and poor parental mental health substantially increases the risk of being NEET on transition in early adulthood. These findings reinforce the importance of early family-centred interventions and the need for structural policies that support child development, educational attainment and the transition from school to work.

## Contributors

NKA carried out the statistical analyses and led the drafting of the manuscript with KU and LA. NKA, LA, KU and DTR contributed to the study design and analysis plan. All authors contributed to the conception of the study, interpretation of results and critically reviewed the manuscript for its intellectual content. NKA and LA accessed and verified the underlying data, and all authors had access. All authors approved the final manuscript as submitted and agree to be accountable for all aspects of the work.

## Patient and public Involvement

Patients and/or the public were not involved in the design, or conduct, or reporting, or dissemination plans of this research.

## Data availability statement

The data that support the findings of this study are available from the UK Data Service by application, under license. For further information on how to obtain the dataset, visit the UK Data Service website (https://ukdataservice.ac.uk/).

## Ethic Statements

### Patient Consent for Publication

Not applicable

### Ethics approval

The data collection of MCS is approved by the UK National Health Service Research Ethics Committee, and written consent was obtained from all participating parents at each survey; MCS1: South West MREC (MREC/01/6/19); MCS2 and MCS3: London MREC (MREC/03/2/022, 05/MRE02/46); MCS4: Yorkshire MREC (07/MRE03/32); MCS5: Yorkshire and The Humber-Leeds East (11/YH/0203); MCS6: London MREC(13/LO/1786). No additional ethical approval was needed for this secondary data analysis.

## References

1. Rahmani H, Groot W, Rahmani AM. Unravelling the NEET phenomenon: a systematic literature review and meta-analysis of risk factors for youth not in education, employment, or training. International Journal of Adolescence and Youth. 2024;29(1). doi: 10.1080/02673843.2024.2331576.

2. Andy P. NEET: Young People Not in Education, Employment or Training (Research Briefing). House of Commons Library, 2021.

3. Eurofound. NEETs–Young people not in employment, education or training: Characteristics, costs and policy responses in Europe. Luxembourg 2012.

4. Lecerf M. NEETs: who are they? Being young and not in employment, education or training today. European Parliamentary Research Service; 2017.

5. Office for National Statistics. Young people not in education, employment or training (NEET), UK: February 2025. ONS website, statistical bulletin; 2025.

6. Feng Z, Ralston K, Everington D, Dibben C. Long term health effects of NEET experiences: evidence from a longitudinal analysis of young people in Scotland. International Journal of Population Data Science. 2017;1(1):327. doi: 10.23889/ijpds.v1i1.327. PubMed Central PMCID: PMCPMC9351037.

7. Gariepy G, Danna SM, Hawke L, Henderson J, Iyer SN. The mental health of young people who are not in education, employment, or training: a systematic review and meta-analysis. Soc Psychiatry Psychiatr Epidemiol. 2022;57(6):1107–21. Epub 20211221. doi: 10.1007/s00127-021-02212-8. PubMed PMID: 34931257; PubMed Central PMCID: PMCPMC8687877.

8. Doring N, Lundberg M, Dalman C, Hemmingsson T, Rasmussen F, Wallin AS, et al. Labour market position of young people and premature mortality in adult life: A 26-year follow-up of 569 528 Swedish 18 year-olds. Lancet Reg Health Eur. 2021;3:100048. Epub 20210211. doi: 10.1016/j.lanepe.2021.100048. PubMed PMID: 34557798; PubMed Central PMCID: PMCPMC8454531.

9. Chandler RF, Santos Lozada AR. Health status among NEET adolescents and young adults in the United States, 2016-2018. SSM Popul Health. 2021;14:100814. Epub 20210506. doi: 10.1016/j.ssmph.2021.100814. PubMed PMID: 34027012; PubMed Central PMCID: PMCPMC8134726.

10. Thompson R. Individualisation and social exclusion: the case of young people not in education, employment or training. Oxford Review of Education. 2011;37(6):785–802. doi: 10.1080/03054985.2011.636507.

11. Bell D, Blanchflower DG. Young people and the Great Recession. Oxford Review of Economic Policy. 2011;27(2):241–67. doi: 10.1093/oxrep/grr011.

12. Bell D, Blanchflower DG. UK Unemployment in the Great Recession. National Institute Economic Review. 2010:214.

13. Cabral FoJ. Key drivers of NEET phenomenon among youth people in Senegal. Economics Bulletin. 2018;38(1):248–61.

14. Duckworth K, Schoon I. Beating the Odds: Exploring the Impact of Social Risk on Young People’s School-to-Work Transitions during Recession in the UK. National Institute Economic Review. 2020;222:R38–R51. doi: 10.1177/002795011222200104.

15. Giret J-F, Guégnard C, Joseph O. School-to-work transition in France: the role of education in escaping long-term NEET trajectories. International Journal of Lifelong Education. 2020;39(5-6):428–44. doi: 10.1080/02601370.2020.1796835.

16. Pitkänen J, Remes H, Moustgaard H, Martikainen P. Parental socioeconomic resources and adverse childhood experiences as predictors of not in education, employment, or training: a Finnish register-based longitudinal study. Journal of Youth Studies. 2019;24(1):1–18. doi: 10.1080/13676261.2019.1679745.

17. Szpakowicz D. Problematising engagement with technologies in transitions of young people identified as ‘Not in Education, Employment or Training’ (NEET) in Scotland. Journal of Youth Studies. 2022;26(9):1200–18. doi: 10.1080/13676261.2022.2080538.

18. Basta M, Karakonstantis S, Koutra K, Dafermos V, Papargiris A, Drakaki M, et al. NEET status among young Greeks: Association with mental health and substance use. J Affect Disord. 2019;253:210–7. Epub 20190424. doi: 10.1016/j.jad.2019.04.095. PubMed PMID: 31054446.

19. Benjet C, Hernandez-Montoya D, Borges G, Mendez E, Medina-Mora ME, Aguilar-Gaxiola S. Youth who neither study nor work: mental health, education and employment. Salud Publica Mex. 2012;54(4):410–7. doi: 10.1590/s0036-36342012000400011. PubMed PMID: 22832833.

20. Tayfur SN, Prior S, Roy AS, Maciver D, Forsyth K, Fitzpatrick LI. Associations between Adolescent Psychosocial Factors and Disengagement from Education and Employment in Young Adulthood among Individuals with Common Mental Health Problems. J Youth Adolesc. 2022;51(7):1397–408. Epub 20220311. doi: 10.1007/s10964-022-01592-7. PubMed PMID: 35275309; PubMed Central PMCID: PMCPMC9135777.

21. de Vries TR, Arends I, Oldehinkel AJ, Bultmann U. Associations between type of childhood adversities and labour market participation and employment conditions in young adults. J Epidemiol Community Health. 2023;77(4):230–6. Epub 20230217. doi: 10.1136/jech-2022-219574. PubMed PMID: 36805940; PubMed Central PMCID: PMCPMC10086506.

22. Elsenburg LK, Kreshpaj B, Andersen SH, de Vries TR, Thielen K, Rod NH. Childhood adversity trajectories and not being in education, employment, or training during early adulthood: The Danish life course cohort (DANLIFE). Soc Sci Med. 2025;371:117841. Epub 20250211. doi: 10.1016/j.socscimed.2025.117841. PubMed PMID: 40037152.

23. Smith KE, Pollak SD. Early life stress and development: potential mechanisms for adverse outcomes. J Neurodev Disord. 2020;12(1):34. Epub 20201216. doi: 10.1186/s11689-020-09337-y. PubMed PMID: 33327939; PubMed Central PMCID: PMCPMC7745388.

24. Bennetsen SK, Kreshpaj B, Andersen SH, Rudmer de Vries T, Thielen K, Lange T, et al. Childhood adversity, early school leaving and long-term social benefit use: A longitudinal mediation analysis of a population-wide study. Soc Sci Med. 2025;370:117770. Epub 20250201. doi: 10.1016/j.socscimed.2025.117770. PubMed PMID: 39999576.

25. Kreshpaj B, Elsenburg LK, Andersen SH, De Vries TR, Thielen K, Rod NH. Association between childhood adversity and use of the health, social, and justice systems in Denmark (DANLIFE): a nationwide cohort study. Lancet Public Health. 2025;10(1):e29–e35. Epub 20241213. doi: 10.1016/S2468-2667(24)00242-1. PubMed PMID: 39681123.

26. Adjei NK, Schluter DK, Straatmann VS, Melis G, Fleming KM, McGovern R, et al. Impact of poverty and family adversity on adolescent health: a multi-trajectory analysis using the UK Millennium Cohort Study. Lancet Reg Health Eur. 2022;13:100279. Epub 20211130. doi: 10.1016/j.lanepe.2021.100279. PubMed PMID: 35199082; PubMed Central PMCID: PMCPMC8841277.

27. Alwin DF. Integrating varieties of life course concepts. J Gerontol B Psychol Sci Soc Sci. 2012;67(2):206–20. Epub 20120307. doi: 10.1093/geronb/gbr146. PubMed PMID: 22399576; PubMed Central PMCID: PMCPMC3307990.

28. Nagin DS, Jones BL, Passos VL, Tremblay RE. Group-based multi-trajectory modeling. Stat Methods Med Res. 2018;27(7):2015–23. Epub 20161017. doi: 10.1177/0962280216673085. PubMed PMID: 29846144.

29. Nagin DS, Odgers CL. Group-based trajectory modeling in clinical research. Annu Rev Clin Psychol. 2010;6(Volume 6, 2010):109–38. doi: 10.1146/annurev.clinpsy.121208.131413. PubMed PMID: 20192788.

30. Taylor-Robinson DC, Straatmann VS, Whitehead M. Adverse childhood experiences or adverse childhood socioeconomic conditions? Lancet Public Health. 2018;3(6):e262–e3. doi: 10.1016/S2468-2667(18)30094-X. PubMed PMID: 29776799.

31. Connelly R, Platt L. Cohort profile: UK Millennium Cohort Study (MCS). Int J Epidemiol. 2014;43(6):1719–25. Epub 20140217. doi: 10.1093/ije/dyu001. PubMed PMID: 24550246.

32. Izquierdo Llanes G, Salcedo Galiano A. Why does equivalization matter? An application to the monetary poverty in the sustainable development goals framework. Quality & Quantity. 2023;57(3):2575–89.

33. Office for National Statistics. Young people not in education, employment or training (NEET), UK. 2025.

34. Lacey RE, Howe LD, Kelly-Irving M, Bartley M, Kelly Y. The Clustering of Adverse Childhood Experiences in the Avon Longitudinal Study of Parents and Children: Are Gender and Poverty Important? J Interpers Violence. 2022;37(5-6):2218–41. Epub 20200708. doi: 10.1177/0886260520935096. PubMed PMID: 32639853; PubMed Central PMCID: PMCPMC8918866.

35. Naja H Rod JB, Esben Budtz-Jørgensen, Clara Clipet-Jensen, David Taylor-Robinson, Anne-Marie Nybo Andersen, Nadya Dich, Andreas Rieckmann. Trajectories of childhood adversity and mortality in early adulthood: a population-based cohort study. The Lancet. 2020;396(10249):489–97.

36. Adjei NK, Schluter DK, Melis G, Straatmann VS, Fleming KM, Wickham S, et al. Impact of Parental Mental Health and Poverty on the Health of the Next Generation: A Multi-Trajectory Analysis Using the UK Millennium Cohort Study. J Adolesc Health. 2024;74(1):60–70. Epub 20231011. doi: 10.1016/j.jadohealth.2023.07.029. PubMed PMID: 37831048.

37. Costello EJ, Compton SN, Keeler G, Angold A. Relationships between poverty and psychopathology: a natural experiment. JAMA. 2003;290(15):2023–9. doi: 10.1001/jama.290.15.2023. PubMed PMID: 14559956.

38. GOV.UK. Alternative provision for primary-age pupils in England: a long-term ‘destination’ or a ‘temporary solution’? Research and analysis. GOV.UK, 2022 Contract No.: 21:05:2025.

39. Jones JH, Call TA, Wolford SN, McWey LM. Parental Stress and Child Outcomes: The Mediating Role of Family Conflict. Journal of Child and Family Studies. 2021;30(3):746–56. doi: 10.1007/s10826-021-01904-8.

40. Connell E, Warburton M, Wood M, al e. School absence and Not in Education, Employment or Training. University of Leeds, 2024.

41. Office for National Statistics. How do childhood circumstances affect your chances of poverty as an adult? Office for National Statistics, 2016 Contract No.: 21:05:2025.

42. Office for National Statistics. Intergenerational transmission of disadvantage in the UK & EU. Office for National Statistics, 2014 Contract No.: 21:05:2025.

43. Fewell Z, Davey Smith G, Sterne JA. The impact of residual and unmeasured confounding in epidemiologic studies: a simulation study. Am J Epidemiol. 2007;166(6):646–55. Epub 20070705. doi: 10.1093/aje/kwm165. PubMed PMID: 17615092.

44. Linden A, Mathur MB, VanderWeele TJ. Conducting sensitivity analysis for unmeasured confounding in observational studies using E-values: the evalue package. The Stata Journal. 2020;20(1):162–75.

45. Counil E. Contribution of causal factors to disease burden: how to interpret attributable fractions. Breathe. 2022;17(4).

46. Gartland D, Riggs E, Muyeen S, Giallo R, Afifi TO, MacMillan H, et al. What factors are associated with resilient outcomes in children exposed to social adversity? A systematic review. BMJ Open. 2019;9(4):e024870. Epub 20190411. doi: 10.1136/bmjopen-2018-024870. PubMed PMID: 30975671; PubMed Central PMCID: PMCPMC6500354.

47. Office for National Statistics. Census 2021. 2024.

48. UK Government. The Green Book: appraisal and evaluation in central government. HM Treasury. 2022.

49. Banks J, Karjalainen H, Waters T. Inequalities in disability. Oxford Open Economics. 2024;3(Supplement_1):i529-i48. doi: 10.1093/ooec/odad091.

50. Dunkley E, McGough K. Number of young people not in work or education hits 11-year high. BBC. 2025.

51. McCurdy C, Murphy L. We’ve only just begun: Action to improve young people’s mental health, education and employment. Resolution Foundation; 2024.

52. UK Government. Tackling Child Poverty: Developing Our Strategy (Policy paper). GOV.UK, 2024 Contract No.: 21:05:2025.

53. Marmot M. Health equity in England: the Marmot review 10 years on. BMJ. 2020;368:m693. Epub 20200224. doi: 10.1136/bmj.m693. PubMed PMID: 32094110.

54. Department for Work and Pensions. Thousands of young people set to benefit from new support into work and training. GOV.UK; 2025.

