## Supplementary Appendix for "Poverty and family adversity trajectories and Not in Education, Employment or Training (NEET) status in early adulthood: Evidence from the UK Millennium Cohort Study"

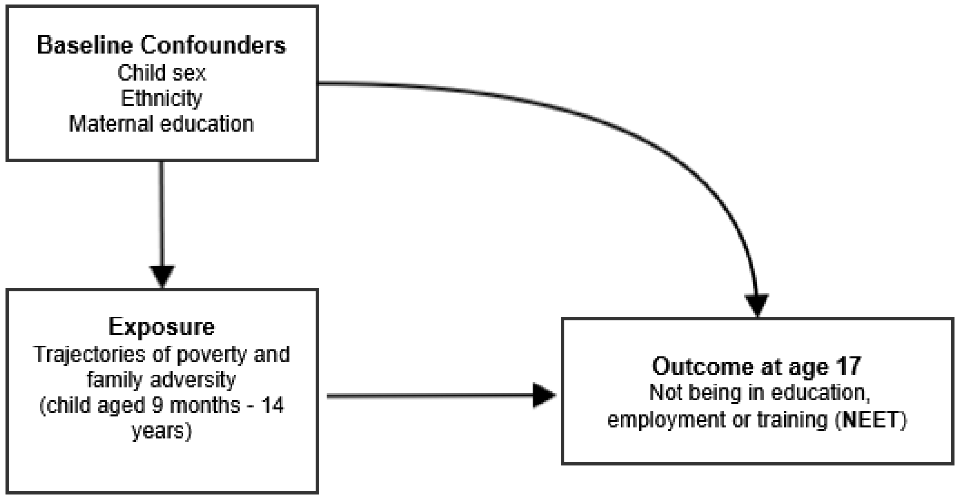


**Figure S1**. Directed acyclic graph for the current study

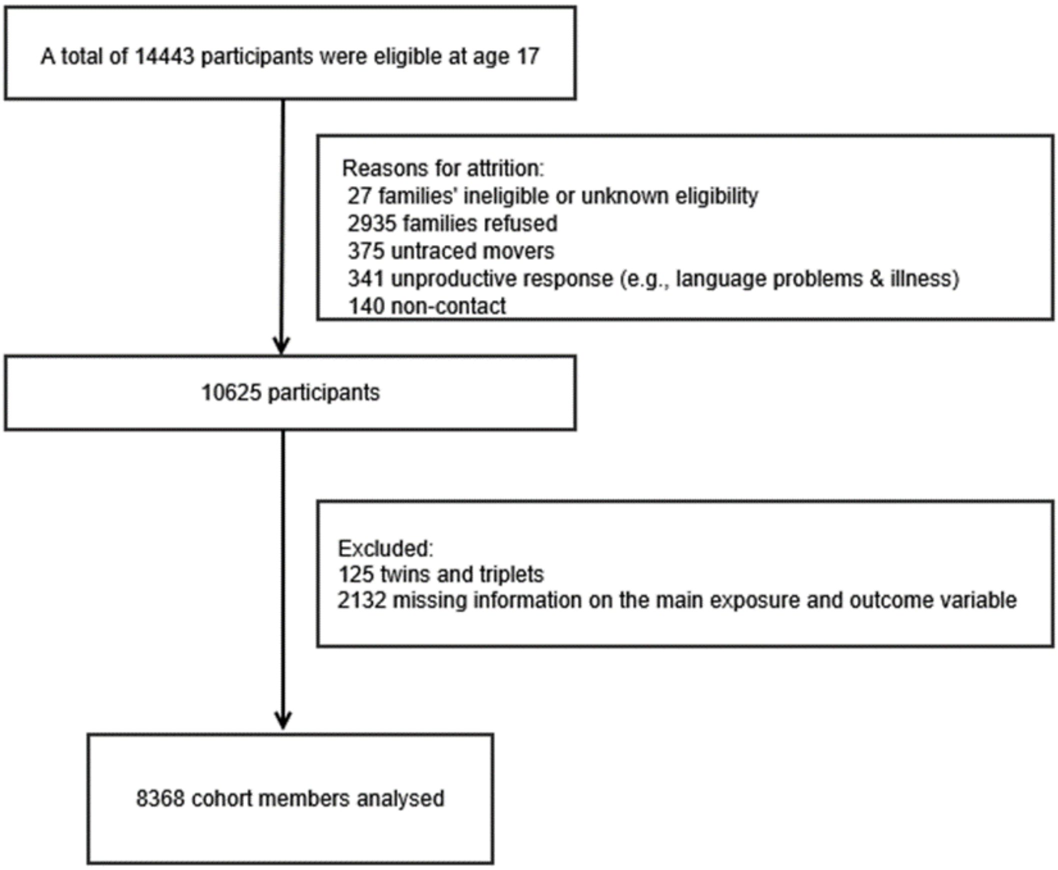


**Figure S2**. Study flow diagram showing inclusion and exclusion of cohort participant.

**Box S1.** Description of measurements assessed for trajectory exposures

- **Parental mental ill health (Child aged 9 months)** – Rutter Malaise Inventory (RMI) scale was used to assess parental mental ill health· A shortened 9-item self-completed version of the RMI measuring depression, anxiety and psychosomatic illness was used· The 9-item short form included items 'feel tired most of the time', 'feel miserable or depressed', 'worried about things', 'often get into violent rage' 'suddenly become scared for no good reason', 'easily upset or irritated', 'constantly keyed up or jittery', 'every little thing gets on nerves and wears you out', and 'heart race like mad'· Scores from these items were summed, and we used a validated cut off for mental ill health ['yes (scores >=4)/no']·

- **Parental mental ill health (Child aged 3 to 14 years) –** Kessler 6 (K6) scale was used to assess parental mental ill health in the last 30 days asking the responders how often they felt depressed, hopeless, restless or fidgety, worthless, or that everything was an effort· Respondents answered on a five-point scale from 1(all the time) to 5 (none of the time)· We reversed and rescaled all items from 0 to 4 for analysis purposes, so that high scores indicate high levels of psychological distress· We used a validated cutoff widely used in previous studies ['yes (scores >=6)/no']

- **Frequent parental alcohol use (Child aged 9 months to 7 years) –** the main responder answered a question about their usual frequency of alcohol consumption ('*Every day, 5-6 times per week, 3-4 times per week, 1-2 per week, 1-2 per month, less than once a month or never'*)·
  Dichotomised: [every day and 5-6 times per week (Yes) vs· 3-4 per week/1-2 per week/ 1-2 per month/never (No)]

- **Frequent parental alcohol use (Child aged 11 to 14 years) –** the main responder answered a question about the usual frequency of alcohol consumption *('>=4 times per week, 2-3 times per week, 2-4 times per month, monthly or less, or never')*·
  Dichotomised: [4 or more times a week (Yes) vs· 2-3 per week/2-4 per month/ monthly or less/never (No)]

- **Domestic violence and abuse (Child aged 9 months to 14 years) –** the main responder was asked about the use of physical force by the partner in relationship ('*Yes, No'*)

- **Poverty (Child aged 9 months to 14 years)** – relative income poverty, defined as household equivalised income of less than 60% of national median household income equivalised according to the Organisation for Economic Co-operation and Development (OECD) household equivalence scale

**Technical Description S1.** Population Attributable Fraction (PAF)

We calculated the PAF for exposure to any trajectory compared to the low poverty and family adversity trajectory group according to the general formula:

$$PAF = \sum_{i=0}^{n} P_i \left( \frac{RR_i - 1}{RR_i} \right)$$

where $P_i$ represents the proportion of cases in each exposure (i.e., each trajectory group) and $RR_i$ represents the adjusted RR of NEET for each trajectory group compared to the low poverty and family adversity group.

The PAF is calculated comparing two scenarios: scenario 1 (a hypothetical scenario in which all children were in the low poverty and adversity trajectory) with scenario 0 (the real world in which there are children in the low poverty and adversity and other trajectories). The reference group is the low poverty and family adversity trajectory.

**Table S1.** Baseline characteristics and outcome by the six estimated trajectory groups, imputed data

| | Family adversity and poverty trajectories | | | | | |
|---|---|---|---|---|---|---|
| **Characteristics** | Low poverty and adversity (3846) | Persistent parental alcohol use (n=706) | Persistent domestic violence and abuse (n=300) | Persistent poor parental mental health (n=991) | Persistent poverty (n=1714) | Persistent poverty and poor parental mental health (n=811) |
| **NEET** | 63 (1.6%) | 9 (1.3%) | 10 (3.3%) | 35 (3.5%) | 95 (5.5%) | 77 (9.5%) |
| **No NEET** | 3783 (98. 4%) | 697 (98.7%) | 290 (96.7%) | 956 (96.5%) | 1619 (95.5%) | 734 (90.5%) |
| **Child's sex** | | | | | | |
| Boy | 1858 (48.3%) | 329 (46.6%) | 154 (50.3%) | 475 (47.9%) | 748 (43.6%) | 422 (52.1%) |
| Girl | 1988 (51.7%) | 377 (53.4%) | 146 (48.7%) | 516 (52.1%) | 966 (56.4%) | 389 (47.9%) |
| **Maternal education** | | | | | | |
| Degree plus | 1212 (31.5%) | 316 (44.8%) | 70 (23.3%) | 216 (21.8%) | 51 (2.9%) | 14 (1.7%) |
| Diploma | 513 (13.3%) | 84 (11.9%) | 48 (16.0%) | 92 (9.3%) | 57 (3.3%) | 16 (2.0%) |
| A-levels | 518 (13.5%) | 71 (10.1%) | 40 (13.3%) | 121 (12.2%) | 115 (6.7%) | 38 (4.6%) |
| GCSE A-C | 1187 (30.9%) | 165 (23.4%) | 90 (30.0%) | 359 (36.2%) | 543 (31.7%) | 258 (31.8%) |
| GCSE D-G | 199 (5.2%) | 27 (3.8%) | 27 (9.0%) | 93 (9.3%) | 235 (13.7%) | 117 (14.4%) |
| None | 217 (5.6%) | 43 (6.1%) | 25 (8.3%) | 110 (11.1%) | 713 (41.6%) | 368 (45.4%) |
| **Maternal ethnicity** | | | | | | |
| White | 3527 (91.7%) | 686 (97.2%) | 255 (85.0%) | 841 (84.7%) | 1067 (62.3%) | 495 (61.0%) |
| Mixed | 20 (0.5%) | 5 (0.7%) | 6 (2.0%) | 7 (0.7%) | 27 (1.7%) | 138 (2.2%) |
| Indian | 121 (3.2%) | 3 (0.4%) | 17 (5.7%) | 44 (4.4%) | 54 (3.1%) | 20 (2.5%) |
| Pakistani and Bangladeshi | 44 (1.1%) | 1 (0.1%) | 6 (2.0%) | 31 (3.1%) | 410 (23.9%) | 215 (26.5%) |
| Black or Black British | 82 (2.1%) | 4 (0.6%) | 12 (4.0%) | 29 (2.9%) | 111 (6.5%) | 39 (4.8%) |
| Other ethnic groups | 52 (1.4%) | 7 (1.0%) | 4 (1.3%) | 39 (3.9%) | 43 (2.5%) | 24 (3.0%) |

**Table S2.** Associations of poverty and family adversity trajectories and being NEET at age 17 years in the UK Millennium cohort study, unadjusted model

| Variables | NEET |
|---|---|
| **Trajectories** | |
| Low poverty and adversity | **Ref.** |
| Persistent parental alcohol use | 0.77 (0.38-1.56) |
| Persistent domestic violence and abuse | 2.07 (1.05-4.07) |
| Persistent poor parental mental health | 2.20 (1.44-3.34) |
| Persistent poverty | 3.52 (2.54-4.87) |
| Persistent poverty and poor parental mental health | 6.30 (4.47-8.86) |

**Table S3.** Associations of poverty and family adversity trajectories and being NEET at age 17 years in the UK Millennium cohort study, imputed data

| Variables | NEET |
|---|---|
| **Trajectories** | |
| Low poverty and adversity | **Ref.** |
| Persistent parental alcohol use | 0.76 (0.37-1.53) |
| Persistent domestic violence and abuse | 2.10 (1.06-4.14) |
| Persistent poor parental mental health | 2.16 (1.41-3.29) |
| Persistent poverty | 3.58 (2.50-5.11) |
| Persistent poverty and poor parental mental health | 6.29 (4.29-9.20) |

Note: model adjusted for child's sex, maternal education and maternal ethnicity

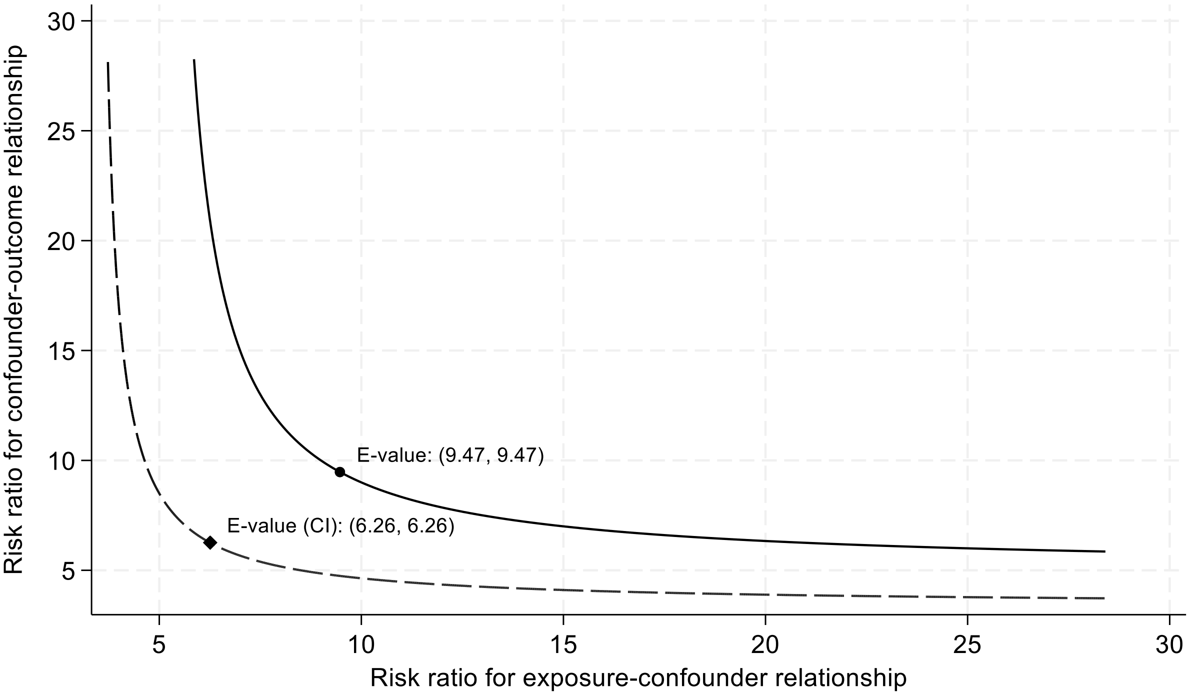


**Figure S3.** Sensitivity analysis for unmeasured confounding using E-values

The E-value is 9.47 with a lower confidence limit (LCL) of 6.26. This means the unmeasured confounder would have to be associated with both family adversity (i.e., poverty and poor parental mental health) and NEET at age 17 by a risk ratio of 9.5 times each. To move the LCL to include no effect, association of an unmeasured confounder would need to be 6.3 or above.
